# Oral hygiene practice, status, and periodontitis among dental outpatient elderly in tertiary hospitals: a cross-sectional study in Dhaka, Bangladesh

**DOI:** 10.1101/2024.10.12.24315397

**Authors:** Sadia Mehjabeen Kanta, Sifat Sharmin, Golam Sharower, Zaki Farhana, Shuvojit Kumar Kundu, Sadhan Kumar Das, Anton Abdulbasah Kamil, Mohammad Meshbahur Rahman

## Abstract

**Background:** Oral health significantly affects older people’s quality of life, overall health, and well-being. The study aimed to assess oral hygiene practice, status, and periodontitis among elderly patients.

**Methods:** A descriptive cross-sectional study was conducted among elderly patients attending dental outpatient departments of tertiary hospitals in Dhaka city from January to December 2022. Data from two hundred and twenty-seven elderly patients were collected by face-to-face interview through a pretested, semi-structured questionnaire and oral examination using the Oral Hygiene Index Simplified (OHI-S), and Community Periodontal Index component of CPITN. Statistics were presented in frequency, mean, percentage, chi-square test, fisher’s exact test, and binary logistic regression.

**Results:** Among 227 elderly patients, the mean (± SD) age was 55.75 (±8.20) years; 51% were female, and 49% were male. Among respondents, 70.48% cleaned their teeth regularly, 61.23% cleaned their teeth once daily, 70.5% used toothbrushes and toothpaste for cleaning their teeth, and only 2.2% used interdental aids. More than half of the respondents, 54%, had poor oral hygiene status, and 55.5% had periodontitis. A significant association existed between periodontitis, age, gender, educational status, occupation, and monthly family income. Older adults aged 66 years and above were 7.2 times more likely to have periodontitis (OR_adj_: 12.637, 95% CI: 3.371-47.371, p < 0.0001). Respondents who engaged in labour work were more likely to have periodontitis than the service holders (OR_adj_:5.855, 95% CI: .594-57.75, p < 0.130). Periodontitis was significantly associated with oral hygiene practices and oral hygiene status. Elderly who did not clean their teeth regularly were roughly 15 times more likely to develop periodontitis (OR _adj_: 9.474, 95% CI: 3.636- 24.689, p < 0.0001). Older adults who did not use toothpaste and toothbrush were 5.1 times more likely to develop periodontitis (OR_adj_: 3.792, 95% CI: 1.758- 8.178, p < 0.001).

**Conclusion:** More than half of the elderly individuals in the study had periodontitis and poor oral hygiene. The research found that the increase in periodontitis was linked to respondents’ socio-demographic characteristics and oral hygiene practices. These findings underscore the importance of maintaining good oral hygiene and getting regular dental checkups for older adults.

## Introduction

Bangladesh is currently experiencing an epidemiological transition characterized by a significant decline in mortality rates from acute, infectious, and parasitic diseases [1–6]. The acceleration of population ageing and an increase in the prevalence of oral health problems and non-communicable diseases will considerably challenge health and social policy planners [7–10].

The World Health Organization (WHO) identified the oral health of older adults as one of its primary concerns due to the growing population of elderly people [11]. Poor oral health in elderly persons is prevalent around the world, with high rates of tooth loss, dental caries, and the prevalence of periodontal disease, xerostomia, and oral precancer/cancer being particularly noticeable [12–14]. Individuals’ physical and psychological well-being, as well as their appearance, pronunciation, ability to chew, taste of food, and sociability, are all impacted by their oral health [15]. In addition to affecting diet, nutrition, and phonetics, poor oral health and tooth loss also endanger general health. Numerous systemic disorders, including cardiovascular disease, bacterial pneumonia, diabetes mellitus, and low birth weight, are affected by oral infection, particularly periodontitis, on their course and aetiology [16]. Thus, good oral health is essential for improved systemic health and healthy dental status.

Proper oral hygiene is the cornerstone of good oral health, preventing eighty percent of oral health problems [17]. Effective oral hygiene and plaque removal are the most crucial methods in preventing periodontal disease since dental plaque is the principal etiological factor in the pathogenesis of this disease [18]. Periodontal disease is a group of inflammatory conditions affecting the gingiva, bone, and periodontal ligament, which can result in tooth loss and spread inflammation throughout the body [19]. Gingivitis, a reversible inflammation, is the first stage of periodontal disease, which can progress to periodontitis, an irreversible infection [20]. One-third of the general population suffers from periodontitis, which is more prevalent in older persons than in younger and middle-aged people [21].

A patient’s dietary habits, quality of life, and sense of self-worth are all impacted by severe periodontal disease, which also raises the chance of multiple tooth loss, masticatory dysfunction, and edentulousness. Beyond its effects on the orofacial structures, severe periodontal disease has been associated with several systemic conditions, including diabetes mellitus, cardiovascular disease, respiratory illnesses, renal disorders, and an elevated risk of head and neck cancers [22]. Therefore, it is imperative to address periodontitis in the elderly population. Nevertheless, there is a shortage of information on the oral health of older adults in Bangladesh [23]. Therefore, it is vital to identify the extent of oral hygiene practice and assess the periodontal health status among older adults. Furthermore, the study’s findings may help the responsible stakeholders take necessary interventions to raise healthier oral hygiene and oral health status among older adults concerning public health activities.

## Methods

### Ethical consent and permission for data collection

This study was approved by the institutional review board of the National Institute of Preventive and Social Medicine (NIPSOM), Bangladesh (Ref No: NIPSOM/IRB/2017/09). The declaration of Helsinki (revised version) was followed to conduct the study. Both written and verbal consent was obtained before initiating the interview. A brief on the aims and objectives was given to the participants. Participants who agreed to give consent were finally included in the study.

### Study setting and participants

The study was a descriptive cross-sectional study conducted among elderly patients who attended the dental outpatient departments of selected hospitals in Dhaka city within one year, from January to December 2022. The elderly patients aged 46years and above attending dental OPD of the selected hospitals (1. Shaheed Suhrawardy Medical College & Hospital, Sher-E-Bangla Nagar, Dhaka 2. Dhaka Dental College Hospital, Mirpur-14, Dhaka 3. Probin Hospital, Agargoan, Dhaka) were included in our study.

### Sampling technique and sample size

A convenient type of non-probability sampling technique was followed in the study. The sample size of the study was calculated by using the formula below:

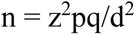

Here, n=assumed/desired sample size; z=the standard normal deviation, usually set at 1.96 at a 95% confidence level; p=proportion of periodontitis among elderly people is 82%=0.82, obtained by the literature search [24]; q=1−p; and d=Margin of error (5%) = 0.05.

The sample size for the study is

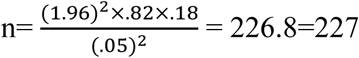

Data were collected from 227 patients after obtaining informed written consent.

### Selection criteria

The inclusion criteria were: (1) Elderly patients aged 46 years and above attending dental OPD (2) Willing to participate and given informed written consent. And the exclusion criteria were Patients with mental or physical disabilities, those who were edentulous, and those who did not have at least two teeth in each sextant were all excluded from this study.

### Data collection tool and procedures

A pretested semi-structured questionnaire was used to collect the data through face-to-face interviews. The questionnaire contained a checklist for oral examination. The simplified Oral Hygiene Index (OHI-S) was used to assess individuals’ oral hygiene status. The Community periodontal index (CPI) component of the Community Periodontal Index of Treatment Need (CPITN) was used to measure the periodontal disease condition of the respondents. Oral examinations were performed in the outpatient department of the hospital. The individuals to be examined remained seated on a chair, and the examiner stood. The examiner was vested with a mask and gloves and used a WHO community periodontal index probe, mirror, and caries probe (dental explorer).

### Simplified Oral Hygiene Index (OHI-S)

A simplified oral hygiene index with debris (plaque) and calculus components was first described by J. Greene and J.R. Vermillion in 1964 [25]. OHI-S is an index that measures the current oral hygiene status based on the number of debris and calculus on six tooth surfaces in the mouth. Six tooth surfaces were scored, four posterior and two anterior. The index values were calculated from the recordings of the calculus and debris scores. The debris scores were added and divided by the number of surfaces scored for each person. A score for a group of individuals was obtained, and the average of the individual scores was calculated. The average group or individual score was defined as the Simplified Debris Index (DI-S). The same process was used to obtain the calculus scores or the Simplified Calculus Index (CI-S). To obtain the Simplified Oral Hygiene Index (OHI- S), the average group or individual calculus and debris scores were added. Range of scores for oral hygiene index: Good: 0-1.2, Fair: 1.3-3, Poor: 3.1-6. A clinical examination of plaque and calculus was done using a caries probe (dental explorer) and mouth mirror.

### Community Periodontal Index of Treatment Needs (CPITN)

The Community Periodontal Index of Treatment Needs (CPITN), introduced by the World Health Organization (WHO) in 1977, included periodontal probing on ten index teeth [26]. CPITN uses a periodontal probe to assess the presence or absence of gingival bleeding on probing, supra or sub-gingival calculus, and periodontal pockets. The mouth was divided into six sextants, and the worst score in each sextant was recorded. The 10 index teeth were examined at mesial and distal proximal sites on the buccal and lingual/palatal sides. Periodontal status was evaluated using the Community Periodontal Index (CPI) component of the CPITN index. About periodontitis (CPI ≥3) and non-periodontitis (CPI ≤2), all research variables were examined [24].

The scoring criteria of CPITN are as follows Score 0: Healthy periodontal conditions or no periodontal disease; Score 1: Bleeding on probing; Score 2: Calculus with plaque felt by probing, Score 3: Shallow periodontal pockets (4 to 5 millimeters), Score 4: Deep periodontal pockets (6 millimeters or more). Pocket depths were measured with a graduated WHO CPI probe.

### Data quality control and statistical analysis

To ensure the reliability and validity of the study results, we used several techniques: (1) conducted pretesting of our study and revised the questionnaire (2) checked the data and fixed errors by observing descriptive statistics; (3) employed an updated version of statistical software in data analysis.

In data analysis, descriptive statistics, such as frequency, percentage, mean, and SD (standard deviation), were used to describe the respondents’ socio-demographic characteristics and selective attributes. Inferential statistics including Chi-square test, Fisher’s exact test, and Binary logistic regression were also done. A p-value of 5% was considered significant at the 95% confidence interval. After data collection and processing, this study compiled and analyzed data according to the objectives and variables by IBM Software – SPSS (Statistical Package for Social Science) version-26.

## Result

### Socio-demographic profile of the respondents

Table 1 shows the socio-demographic characteristics of the patients who participated in our study. According to the distribution of respondents by age group, the 46-55-year-old age group had the highest frequency, and the mean age was 55.72 years. Among 227 respondents, 51% were female, and 49% were male. It was observed that 53 (23.36%) respondents had no formal education (Table 1).

**Table 1:** Socio-demographic characteristics of the respondents.

| Variables | f (%) |
| --- | --- |
| **Age group(years)** |  |
| 46-55 | 110 (48.5) |
| 56-65 | 89 (39.2) |
| 66 and above | 28 (12.3) |
| **Gender** |  |
| Male | 111 (49) |
| Female | 116 (51) |
| **Educational Status** |  |
| No formal education | 53 (23.3) |
| Primary | 46 (20.3) |
| Secondary | 63 (27.8) |
| Higher secondary and above | 65 (28.6) |
| **Occupation** |  |
| Service holder | 38 (16.7) |
| Businessman | 24 (10.6) |
| Labourer | 16 (7.0) |
| Homemaker | 108 (47.6) |
| Unemployed/ Retired | 41 (18.1) |
| **Monthly family income** |  |
| 5000-24000 | 146 (64.32) |
| 24001-44000 | 38 (16.74) |
| 44001 and above | 43 (18.9) |

About 46 (20.3%), 63 (27.85%), and 65 (28.6%) had completed primary, secondary, higher secondary and above, respectively. Almost half of the respondents were homemakers, 108 (47.6%), and the percentage of labourer respondents was the lowest (7%). It was also observed that the highest proportion of respondents, 146 (64.32%), had monthly family income within 5000–24000 BDT (Table-1).

### Oral hygiene practices of respondents

It was found that most respondents, 160 (70.48%), cleaned their teeth regularly, and 67 (29.52%) did not clean their teeth regularly. Most of them, 222 (97.8%), did not use interdental aids, 219(96.5%) did not use mouthwash, 200 (88.1%) did not clean their tongue, whereas 59 (26%) respondents rinsed their mouth after a meal. It was found that the majority of the respondents, 222 (97.8%), visited the dentist only when they had a problem. Among 227 respondents, 139 (61.23%) cleaned their teeth once daily, and 88 (38.77%) cleaned their teeth twice or more daily. Most of the respondents, 160 (70.5%), used toothbrushes and toothpaste for cleaning their teeth, whereas 67 (29.5%) used other materials (tooth powder, charcoal, miswak, or chew stick) (Table-2).

**Table 2:**
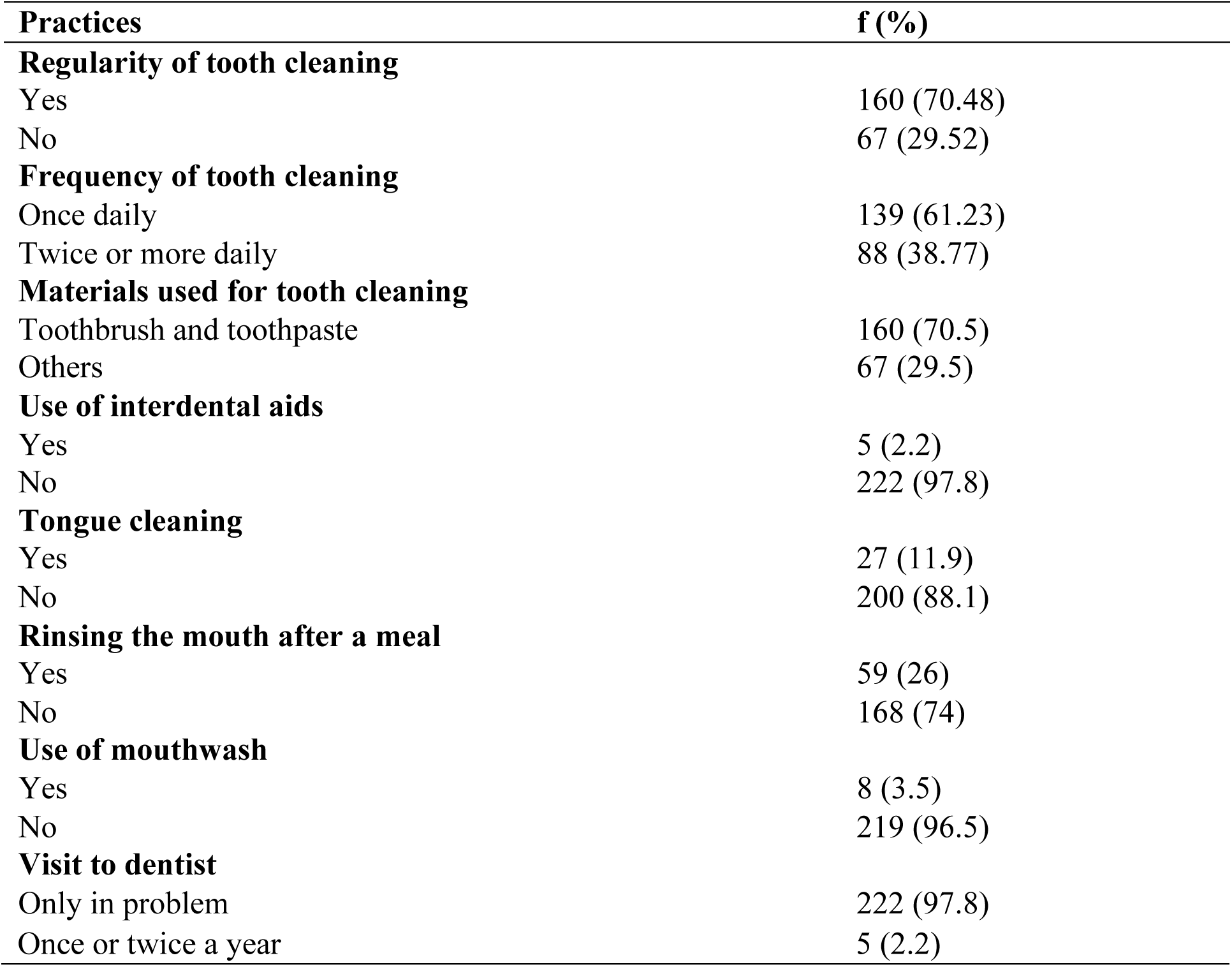
Oral hygiene practices of the respondents.

### Oral Hygiene status, Periodontal Health status, Periodontitis of Respondents of respondents

Figure 1A illustrates the distribution of respondents based on oral hygiene status. Out of 227 participants, 123 (54%) exhibited poor oral hygiene, 69 (30%) demonstrated fair hygiene, and the remaining 35 (16%) had good oral hygiene.

**Figure 1.**
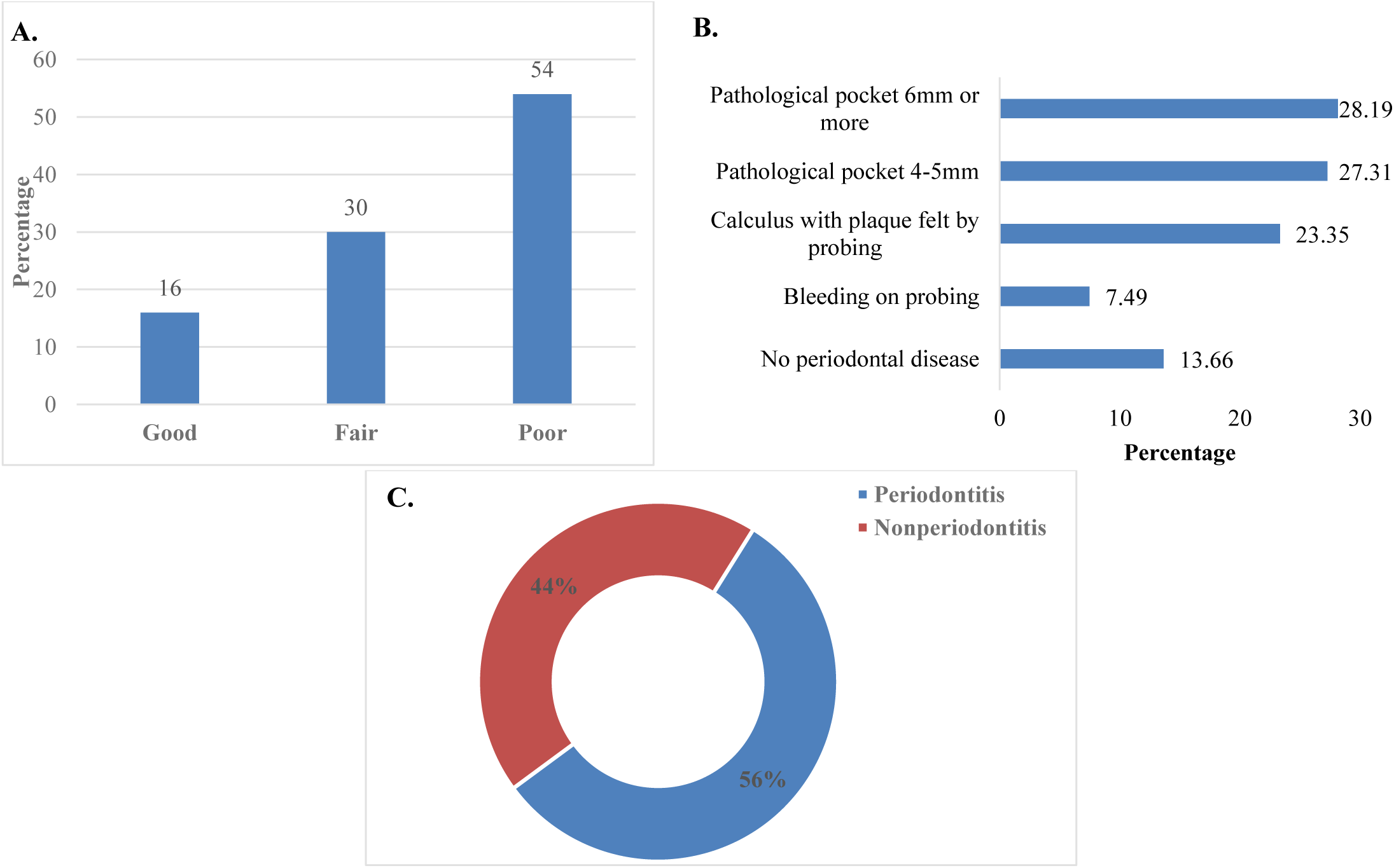
A. Distribution of respondents by oral hygiene status; B. Distribution of respondents by periodontal health status; C. Distribution of respondents by presence of periodontitis.

Figure 1B shows the distribution of respondents by periodontal health status. Of the total, 31 respondents (13.66%) were free from periodontal disease, 17 (7.49%) exhibited bleeding on probing, 53 (23.35%) presented with calculus detectable by probing, 62 (27.31%) had pathological pockets of 4–5 mm, and 64 (28.19%) exhibited pockets measuring 6 mm or more.

Figure 1C depicts the presence of periodontitis among respondents. Out of the 227 individuals, 126 (55.5%) were diagnosed with periodontitis, while 101 (44.5%) did not present with the condition.

### Association of respondent’s socio-demographic characteristics with periodontitis

Table 3 shows the distribution of periodontitis concerning socio-demographic characteristics. Age group, sex, educational status, occupation, and monthly family income were significantly associated with periodontitis.

**Table 3:** Distribution of periodontitis concerning socio-demographic characteristics.

| Variable | Periodontitis<br>f (%) | Non-periodontitis<br>f (%) | P value |
| --- | --- | --- | --- |
| Age group |  |  |  |
| 46-55 years | 50 (45.50) | 60 (54.50) | 0.001 |
| 56-65 years | 52 (58.40) | 37 (41.60) |  |
| 66 years and above | 24 (85.7) | 4 (14.3) |  |
| Gender |  |  |  |
| Male | 69 (62.20) | 42 (37.80) | 0.048 |
| Female | 57 (49.10) | 59 (50.90) |  |
| Educational Status |  |  |  |
| No formal education | 45 (84.90) | 8 (15.10) | 0.0001 |
| Primary | 31 (67.40) | 15 (32.60) |  |
| Secondary | 26 (41.30) | 37 (58.70) |  |
| Higher Secondary and above | 24 (36.9) | 41 (63.1) |  |
| Occupation |  |  |  |
| Service holder | 17 (44.70) | 21 (55.30) | 0.019 |
| Businessman | 13 (54.20) | 11 (45.80) |  |
| Laborer | 15 (93.80) | 1 (6.30) |  |
| Homemaker | 57 (52.80) | 51 (47.20) |  |
| Unemployed/ Retired | 24 (58.6) | 17 (41.5) |  |
| Monthly family income (in taka) |  |  |  |
| 5000-24000 | 95 (65.10) | 51 (34.90) | 0.0001 |
| 24001-44000 | 16 (42.10) | 22 (57.90) |  |
| 44001 and above | 15 (34.9) | 28 (65.1) |  |

### Association of respondent’s oral hygiene practices, status with periodontitis

Table 4 shows the distribution of periodontitis regarding oral hygiene practices and status. Regularity, frequency, materials used for tooth cleaning, use of interdental aids, tongue cleaning, rinsing mouth after meal, use of mouthwash, and visits to the dentist were significantly associated with periodontitis. Regarding oral hygiene status, it was found that 100% (35) of respondents with good oral hygiene status had not periodontitis, and 97.60% (120) of respondents with poor oral hygiene status had periodontitis. The association was found statistically significant (p <0.05).

**Table 4:** Association between oral hygiene practices, status, and periodontitis.

| Variable | Periodontitis<br>f (%) | Non-periodontitis<br>f (%) | P value |
| --- | --- | --- | --- |
| Regularity of tooth cleaning |  |  |  |
| Yes | 65 (40.60) | 95 (59.40) | 0.0001 |
| No | 61 (91) | 6 (9) |  |
| Frequency of tooth cleaning |  |  |  |
| Once | 98 (70.50) | 41 (29.50) | 0.0001 |
| Twice or more daily | 28 (31.80) | 60 (68.20) |  |
| Materials used for tooth cleaning |  |  |  |
| Toothbrush and toothpaste | 72 (45.00) | 88 (55.00) | 0.0001 |
| Others | 54 (80.6) | 13 (19.4) |  |
| Use of interdental aids |  |  |  |
| Yes | 0 (0) | 5 (100) | 0.016 |
| No | 126 (56.80) | 96 (43.20) |  |
| Tongue Cleaning |  |  |  |
| Yes | 7 (25.90) | 20 (74.10) | 0.001 |
| No | 119 (59.50) | 81 (40.50) |  |
| Rinsing the mouth after meal |  |  |  |
| Yes | 17 (28.80) | 42 (71.20) | 0.0001 |
| No | 109 (64.90) | 59 (35.10) |  |
| Use of mouthwash |  |  |  |
| Yes | 0 (0.00) | 8 (100.00) | 0.001 |
| No | 126 (57.50) | 93 (42.50) |  |
| **Visit to dentist** |  |  |  |
| Only in problems | 126 (56.8) | 96 (43.2) | 0.012 |
| Once or twice a year | 0 (0.00) | 5 (100.00) |  |
| **Oral hygiene status** |  |  |  |
| Good | 0 (0) | 35 (100) | 0.0001 |
| Fair | 6 (8.7) | 63 (91.3) |  |
| Poor | 120 (97.60) | 3 (2.40) |  |

### Associated factors of periodontitis

Socio-demographic characteristics associated with periodontitis were identified by unadjusted odds ratio and adjusted odds ratio with 95% confidence intervals after adjusting for several important covariates (Table 5). Older adults aged 66 years and above were 7.2times more likely to have periodontitis than the 46-55years age group (OR_un-adj_: 7.2, 95% CI: 2.342-22.135, p < 0.001; OR _adj_: 12.637, 95% CI: 3.371-47.371, p < 0.0001). Male older adults were more likely to develop periodontitis than female older adults (OR_un-adj_: 1.7, 95% CI: 1.002-2.886, p < 0.049; OR _adj_: 3.309, 95% CI: .859-12.747, p < 0.082). Education had a significant impact on periodontitis. Older adults having higher secondary or above education had less chance of developing periodontitis (OR_un-adj_: .104, 95% CI: .042-.257, p < 0.0001; OR _adj_: .100, 95% CI: .032-.307, p < 0.0001). Regarding the impact of occupation on periodontitis, laborers were more likely to have periodontitis than the service holders (OR_un-adj_: 18.529, 95% CI: 2.218-154.810, p < 0.007; OR _adj_:5.855, 95% CI: .594-57.75, p < 0.130). Monthly family income also significantly impacted periodontitis, where older people had higher incomes (Income. ≥ 44001 BDT) family had a lower chance of developing periodontitis (OR_un-adj_: .288, 95% CI: .141-.587, p < 0.001; OR _adj_: .515, 95% CI: .216-1.227, p < 0.134) (Table 5).

**Table 5:** Multiple logistic regression analysis of the association between periodontitis and socio-demographic characteristics.

| Characteristics | Unadjusted<br>OR | CI | P-value | Adjusted<br>OR | CI | P-value |
| --- | --- | --- | --- | --- | --- | --- |
| **Age** |  |  |  |  |  |  |
| 46-55 | Ref |  |  | Ref |  |  |
| 56-65 | 1.686 | 0.96-2.97 | 0.070 | 1.73 | 0.85-3.53 | 0.13 |
| 66 and above | 7.200 | 2.34-22.14 | 0.001 | 12.64 | 3.37-47.37 | 0.0001 |
| **Gender** |  |  |  |  |  |  |
| Female | Ref |  |  | Ref |  |  |
| Male | 1.701 | 1.00-2.89 | 0.049 | 3.31 | 0.86- 12.75 | 0.082 |
| **Education** |  |  |  |  |  |  |
| No formal<br>education | Ref |  |  | Ref |  |  |
| Primary | 0.367 | 0.14-0.97 | 0.044 | 0.39 | 0.14-1.09 | 0.074 |
| Secondary | 0.125 | 0.05-0.31 | 0.0001 | 0.14 | 0.05-.38 | 0.000 |
| Higher secondary<br>and above | 0.104 | 0.04-0.26 | 0.0001 | 0.10 | 0.03-.31 | 0.0001 |
| **Occupation** |  |  |  |  |  |  |
| Service holder | Ref |  |  | Ref |  |  |
| Businessman | 1.46 | 0.52-4.07 | 0.470 | 1.118 | .332- 3.768 | 0.857 |
| Laborer | 18.53 | 2.22-154.81 | 0.007 | 5.855 | .594-57.75 | 0.130 |
| Homemaker | 1.38 | 0.66-2.90 | 0.395 | 1.788 | .453-7.055 | 0.407 |
| Unemployed/<br>Retired | 1.74 | 0.72-4.25 | 0.221 | 0.747 | .233-2.397 | 0.624 |
| **Monthly family income** |  |  |  |  |  |  |
| 5000-24000 | Ref |  |  | Ref |  |  |
| 24001-44000 | .390 | 0.19-0.81 | 0.011 | 0.502 | 0.20-1.25 | 0.140 |
| 44001 and above | .288 | 0.14-0.59 | 0.001 | 0.515 | 0.22-1.23 | 0.134 |

Regularity of tooth cleaning, frequency of tooth cleaning, the material used for tooth cleaning, and rinsing the mouth after the meal significantly impacted periodontitis (Table 6). Elderly who did not clean their teeth regularly were roughly 15 times more likely to develop periodontitis (OR_un-adj_: 14.85, 95% CI: 6.066- 36.400, p < 0.0001; OR _adj_: 9.474, 95% CI: 3.636- 24.689, p < 0.0001). Similarly, respondents who did not use toothpaste and toothbrush were 5.077 times more likely to develop periodontitis (OR_un-adj_: 5.077, 95% CI: 2.570- 10.030, p < 0.0001; OR _adj_: 3.792, 95% CI: 1.758- 8.178, p < 0.001). Older adults who cleaned their teeth twice or more daily were less likely to develop periodontitis (OR_un-adj_: .195, 95% CI: .000- .348, p < 0.0001; OR _adj_: .403, 95% CI: .216- .856, p < 0.016). Rinsing the mouth after meals also had a significant impact on periodontitis, where older people who did not rinse their mouth after meal had 4.56 times more chance of developing periodontitis (OR_un-adj_: 4.564, 95% CI: 2.391- 8.711, p < 0.002; OR _adj_: 3.14, 95% CI: 1.272- 7.755, p < 0.013) (Table 6).

**Table 6:** Multivariate analysis of the association between periodontitis and Oral Hygiene practices.

| Characteristics | Unadjusted OR | CI | P-value | Adjusted OR | CI | P-value |
| --- | --- | --- | --- | --- | --- | --- |
| **Regularity of tooth cleaning** |  |  |  |  |  |  |
| Yes | Ref |  |  | Ref |  |  |
| No | 14.86 | 6.07- 36.40 | 0.0001 | 9.47 | 3.64- 24.69 | 0.0001 |
| **Frequency of tooth cleaning** |  |  |  |  |  |  |
| Once daily | Ref |  |  | Ref |  |  |
| Twice or more daily | .19 | 0.00- 0.35 | 0.0001 | 0.40 | 0.22-0.86 | 0.016 |
| **Materials used for tooth cleaning** |  |  |  |  |  |  |
| Toothbrush and toothpaste | Ref |  |  | Ref |  |  |
| Others | 5.08 | 2.57- 10.03 | 0.0001 | 3.79 | 1.76- 8.18 | 0.001 |
| **Tongue cleaning** |  |  |  |  |  |  |
| Yes | Ref |  |  | Ref |  |  |
| No | 4.19 | 1.69- 10.38 | 0.002 | 0.83 | 0.23- 3.08 | 0.637 |
| **Rinsing the mouth after meal** |  |  |  |  |  |  |
| Yes | Ref |  |  | Ref |  |  |
| No | 4.56 | 2.39- 8.71 | 0.0001 | 3.14 | 1.27-7.75 | 0.013 |

## Discussion

In developing countries like Bangladesh, where the elderly population is increasing significantly, dental health is especially important but frequently overlooked in terms of an individual’s overall health and well-being. People are more prone to oral health issues as they become older, particularly periodontitis, which can have a serious effect on their general health and quality of life. This study highlighted the lack of oral hygiene practices, increased poor oral hygiene status and the association of periodontitis with several factors.

The present study was conducted among 227 elderly patients attending the dental outpatient department of three hospitals in Dhaka city. Most of the respondents, 110 (48.5%), were aged between 46 and 55 years, which is inconsistent with the study among the older adults in Dhaka, Bangladesh, reporting that among the elderly patients, the highest proportion of the respondents (49.10%) was in the age group of 60-69 years (1). Regarding educational qualification, 63 (27.8%) respondents had completed a secondary educational level, which is the highest frequency, and 53 (23.3%) had no formal education. This result is incompatible with the study among older adults in Bangladesh, which reported that most respondents had no schooling [27]. In this study, 108 (47.6%) of the respondents were homemakers, which is comparable with another study where most were homemakers [28]. According to our study findings, the highest proportion, 146 (64.32%) of respondents, had a monthly family income within 5000 – 24000 Taka, which is in line with the study conducted in the Narayanganj district of Bangladesh, reported the monthly family income of most of the respondents ranged from 4000 to 20000 taka per month [27].

The risk of developing periodontitis was higher in older adults who did not regularly brush their teeth. Brushing teeth once daily is enough to keep your mouth healthy and prevent periodontal and dental cavities [29]. Although the majority of the respondents, 160 (70.48%), cleaned their teeth regularly, the percentage of respondents brushing their teeth twice or more daily was 38.77%, which is similar to a previous study where 151 (90.41%) cleaned their teeth regularly, and 41.91% cleaned their teeth twice daily [27].

Another study conducted in New Delhi, India, revealed that 40 (16.1%) elderly participants brushed twice or more daily, less than the current study’s findings [29]. Notably, 160 (70.5%) respondents used toothbrushes and toothpaste for cleaning teeth. This finding agrees with the previous study, where 79.04% of elderly respondents used toothbrushes for tooth cleaning [27].

The most widely used oral hygiene technique worldwide is tooth brushing, while regular brushing seems less prevalent among older people [30]. Similarly, the results of this study indicate that the risk of developing periodontitis was 5.077 times higher for those who did not use a toothbrush and toothpaste. Chahar *et.al* found that 48.2% of respondents used toothbrushes and toothpaste for tooth cleaning, which is less as compared with the current study [30]. The present study revealed that only 5 (2.2%) used interdental aids such as dental floss and interdental brushes. Likewise, the use of dental floss as an interdental cleaning device was almost negligible in the survey conducted in India [31]. Among the 227 respondents, 27 (11.9%) cleaned their tongue, 200 (88.1%) did not clean their tongue, and 59 (26%) respondents rinsed their mouth after meals, whereas the rest of them did not. These findings are in accordance with the study stating that oral hygiene- awareness and practice among patients in Jodhpur, India, found that only 20% of participants cleaned their tongue either with a toothbrush or tongue cleaner, and only 29% of the sample population rinsed their mouth after eating food [32]. According to this study, older adults who did not rinse their mouths after meals had a 4.56 times increased risk of acquiring periodontitis. Rinsing the mouth after meals also substantially affected the development of periodontitis. Again, most of the respondents, 219(96.5%), did not use mouthwash, which is comparable with a cross-sectional study among older Mexican adults, where 16.5 % of the study participants used mouthwash [33]. The current study also showed that the majority of the respondents, 222 (97.8%), visited the dentist only when they had a problem, consistent with the previous study stating that 54% of the study population visited the dentist only in pain [33].

More than half of the respondents, 123 (54%), had poor oral hygiene status, 69 (30%) had fair, and the rest, 35 (16%), had good oral hygiene status. A cross-sectional study conducted among rural dwellers found that one-third (33.8 %) of the participants had poor oral hygiene status, 57.2% had fair, and only 9% had good oral hygiene status [34]. In comparison, the current study had more participants with poor oral hygiene status. In the study from India [35], among 480 workers, 332(81.4%) respondents had poor oral hygiene status, and only 10 (2.5%) had good oral hygiene status, similar to the present study, though the study population is different. In the current study, it was observed that among 227 respondents, 31 (13.66%) had no periodontal disease, followed by 17 (7.49%) respondents who had bleeding on probing, 53 (23.35%) had calculus with plaque felt by probing, 62 (27.31%) had pathological pocket 4-5mm, 64 (28.19%) had pathological pocket 6mm or more. This finding is comparable with the study among the geriatric population in Delhi, India, which revealed that 25.4 % of older people had healthy periodontium, 71.1% had a periodontal pocket of 6 mm or more, and 2.40% had a pocket depth of 4-5 mm [36].

About 126 (55.5%) were found to have periodontitis, while 101 (44.5%) hadn’t. The study among patients visiting a dental hospital in Dhaka reported that 15% had periodontitis, which is discordant with the present study [37]. Assessment of the association between periodontitis and socio- demographic characteristics of respondents revealed that periodontitis is significantly associated with age group, sex, educational status, occupation, and monthly family income. A previous study [38] found that the prevalence of periodontitis was higher among older participants and those of lower educational attainment, which is in accordance with the current study’s findings. The findings of this study showed that elderly adults with a higher secondary education or above had a lower risk of developing periodontitis. Another study [39] observed that periodontitis was significantly higher in males than females and the old elderly compared to the young elderly, which is consistent with the present study. In the current study, oral hygiene practices were significantly associated with periodontitis. Similarly, earlier research [35] found that periodontal disease was significantly associated with oral hygiene practice. The current study’s result supports another study’s findings [40], suggesting that a lack of regular oral hygiene care was associated with periodontal disease. In contrast, a study among healthcare professionals showed that oral hygiene practices were not associated with periodontitis [41]. This contrast may be due to differences in the study population. A significant association was noticed between oral hygiene status and periodontitis, which supports the finding of a previous study [40] that reported that oral hygiene was significantly associated with periodontal disease. Subjects with the poorest periodontal health also displayed the poorest oral hygiene. Another study [35] found a significant association between periodontitis and oral hygiene status. We must prioritize the core public health functions of evaluation, policy formulation, and assurance. This involves redefining the impact of periodontal disease, assessing the effectiveness of preventive and control measures, and highlighting periodontal disease as a major public health concern[42].

The study provided information about oral hygiene practice, status, and periodontal status of elderly patients attending dental outpatient departments. The findings indicate the need to improve the oral health of older adults through preventive and curative public health measures. Further research with a larger sample size could be done to generalize the results and plan for necessary public health interventions accordingly.

## Conclusion

The findings of the study provide insight into the oral hygiene practice, status, and periodontal health status of elderly patients. A significant association existed between periodontitis, age, gender, educational status, occupation, and monthly family income. Oral hygiene practices and oral hygiene status were found to be significantly associated with periodontitis. These observations underscore the urgent need for targeted interventions to improve oral health education, promote proper oral hygiene practices and awareness regarding maintaining good oral hygiene and regular dental checkups, and enhance access to dental care services for elderly individuals in Bangladesh. Addressing these factors makes mitigating the burden of periodontitis and associated oral health complications possible, thereby enhancing the ageing population’s overall quality of life and well- being.

## Data Availability

The datasets generated and analyzed during the current study are available from the corresponding author on reasonable request to.

## Acknowledgement

We acknowledge the Department of Biostatistics and the Department of Health Education for their technical support during the study. We are also grateful to all participants included in this study.

